# Effect and correlation of intraocular pressure on the refractive status of children and adolescents

**DOI:** 10.1101/2023.05.21.23290157

**Authors:** Nie Yingying, Wu xiaoxiao, Zhang xunlang, Yang Chih-Huang, Song yufeng, Duan Junguo

**Affiliations:** Sichuan Myopia Prevention and Treatment Center of Integrative Medicine, Chengdu University of traditional Chinese Medicine,Ineye Hospital of Chengdu University of Traditional Chinese Medicine,Chengdu,China; Chengdu University of traditional Chinese Medicine; Queensland University of Technology; Wenzhou Medical College

**Keywords:** intraocular pressure, ocular axis, equivalent spherical lens, myopia, children

## Abstract

**Objective:** To explore the effect of IOP on the refractive error in children and adolescents, and to analyze the correlation among IOP, AL and diopter of refraction.

**Methods:** Conducting a cross-sectional study. A total of 3256 students (6511 pairs of eyes) aged between 4 and 15 in Jinniu district who presented for ocular health examination during October 2018 and October 2021 were selected, including 1735 boys (3470 eyes) and 1521 girls (3041 eyes).The differences between groups were analyzed by analysis of variance, correlation analysis by Pearson, data were analyzed by statistical software SPSS 25.0.

**Results:** There were significant differences in SE, IOP and AL between each group (P < 0.001), whereas IOP, AL were positively correlated with SE, respectively. The average IOP of female (17.45 ±2.56mmg) was higher than that of male (17.08 ±2.60mmg), while the mean AL of male (23.46 ±0.81 mm) was longer than that of female (22.91 ±0.83 mm). There was weak or no correlation between IOP and AL (r = 0.126, P < 0.001). No correlation was found between IOP and SE (r = 0.116, P < 0.001). Positive correlation between AL and SE (r = 0.632, P < 0.001) was noted.

**Conclusion:** The increase of myopia degree in children at different ages is manifested by the increase of intraocular pressure and axial length, the increase of axial length is one of the main reasons affecting diopter.However, IOP may not directly lead to increase of myopia in children and adolescents within the range of normal IOP.

**Synopsis:** This study suggests that there is no direct relationship between axial length and refraction or intraocular pressure, and intraocular pressure may not be a direct factor influencing the development of myopia.

---

Clinically, it has been found that the average intraocular pressure (IOP)of patients with myopia is higher than that of normal children. Some scholars suspect that high IOP may promote the growth of ocular axis in children with myopia,however, the influence of IOP level on refractive status is still controversial. By analyzing the influence of IOP on refractive status of students aged 4-15 years in Jinniu District of Chengdu, this paper explored the correlation between IOP and refractive status, which can give us a deeper understanding of the correlation between IOP and myopia.This study suggests that there is no direct relationship between axial length and refraction or IOP, and IOP may not be a direct factor influencing the development of myopia, but we should strengthen the detection of IOP in myopic patients, especially those with high myopia, if necessary, the screening of glaucoma should be excluded and IOP should be reduced for treatment.

Intraocular pressure (IOP), axial length (AL) and diopter of refraction are important research indicators of eye growth and development. The relationship between IOP and refractive status has become a hot research topic today. Clinically, IOP is found to be increased to varying degrees in myopic patients, with the average IOP in myopic patients being higher than the average IOP in normal children^[1-2]^, and the IOP in patients with high myopia being higher than that of low to moderate myopia^[3]^.Some domestic as well as foreign studies ^[4-7]^have concluded that IOP is significantly related to refraction and axial length, and it is well known that the refractive status of children and adolescents is closely related to the length of the ocular axis^[8]^. Myopic patients have a reduced modulus of elasticity of the eye due to the elongation of the wall of the eye, so they may be more susceptible to IOP than farsighted or orthotopic children. Thus, high IOP may contribute to the growth of the eye axis in myopic children. Francis B et al.^[9]^found that postoperative intraocular pressure reduction in glaucoma patients resulted in a certain shortening of the ocular axis. And in an animal study of FDM guinea pigs, drops of an IOP-lowering drug (latanoprost) were found to be effective in slowing the progression of myopia. But this has been questioned by many scholars^[10-12]^, who argue that there is no direct link between IOP and refraction or AL. High myopia patients were confirmed ^[13]^to have 2 to 14 times higher risk of getting glaucoma than myopia, but clinical findings are often associate long axial length, high myopia patients with relatively high intraocular pressure. Therefore, it is particularly important to explore the correlation between IOP, AL and diopter of children as well as adolescents for the prevention and control of myopia and their eye health status.

There are many studies on refraction, AL, IOP and fundus conditions in adults with high myopia. However, few studies are conducted on the correlation of IOP, AL and diopter under different refractive states of hyperopia, emmetropia and low, medium and high myopia in children and adolescents. The focus in the available studies ^[14]^has been on the relationship between IOP and AL in patients with congenital or juvenile glaucoma. In recent years, some experimental animal studies^[15-16]^ have found that increased IOP can cause scleral extension of the eye axis length, thus changing the refractive state. Currently, due to the lack of longitudinal studies, the effect of IOP level on refractive status is still controversial. In this paper, we further discuss the above situation and analyze the effect of IOP on the refractive state of children and adolescents by comparing the IOP of children and adolescents with different refractive errors, which can help investigate the pathogenesis of myopia and explore new ideas for its prevention and control in children and adolescents.

## 1. Objects and methods

### 1.1 Objects

A total of 3256 healthy children and adolescents aged 4-15 years with 6511 eyes were selected. Those with a recent history of keratoplasty lens wear, use of pupil dilators such as atropine and IOP-lowering drugs, etc., cone corneas, suspected fundus diseases and family history of hereditary glaucoma were excluded. And the corrected visual acuity should reach normal standards.

In this study, primary and secondary school students in Jinniu District who went to Ineye Hospital affiliated to Chengdu University of Traditional Chinese Medicine for eye health examination from October 2018 to October 2021 were selected.

### 1.2 grouping

According to their selected baseline spherical equivalent(spherical+ cylindrical /2), the subjects were divided into hyperopic group A (> +0.5D), emmetropic group B (−0.25D ≤ equivalent spherical ≤ +0.50D), low-myopia group C (−0.50D ≥ equivalent spherical ≥ -2.75D), moderate-myopia group D (−3.00D ≥ equivalent spherical ≥ -5.75D), and high-myopia group E (≤-6.00D). The mean and standard deviation of IOP, AL, and SE of the five groups were found, and ANOVA and correlation analysis were performed. Subjects were further divided into two groups according to their gender.

### 1.3 methods

The data of axial length (AL) (unit: mm) was measured by The Sw-9000 optical biometric instrument for the patient examination, and the average value was taken three times. The IOP data (unit: mmHg) was measured by AT555 automatic non-contact IOP, three measurements, data averaged. All patients were treated with medoride (compound tropicamide) for ciliary palsy dilation, 5 min/time, local eye drops three times in a row. Medical optometry was performed on the patient after the light reflection disappeared, including computer optometry, retinoscopy and subjective refraction, to determine the diopter of their eyes. All checks were conducted by experienced optometrist.

### 1.4 Statistical approach

Cross-sectional study, statistical software SPSS25.0 was used for statistical analysis. The quantitative data were linear, homogeneity of variance and follows normal distribution. The data index was expressed as the mean X±S. Subjects were divided into hyperopic group, emmetropic group, low-myopia group, moderate-myopia group and high-myopia group according to equivalent spherical. The difference between groups was analyzed by one-way ANOVA, the multiple comparison between groups was tested by LSD-T test, and the correlation analysis of IOP, AL and SE of subjects were conducted by Pearson. We define P < 0.05 as statistically significant.

## 2. Results

### 2.1 General Information

There were 3,256 children and adolescents aged 4-15 in 6511 eyes, 1735 boys in 3470 eyes and 1521 girls in 3041 eyes. The average age was (7.9±1.4) years, and the equivalent spherical was (+0.35±1.19) D. The average IOP was (17.25±2.59) mmHg and the average AL was (23.20±0.86) mm. There were no statistically significant differences in age, gender, diopter, IOP and AL among the five groups at baseline, as shown in Table 1.

**Table 1.** Comparison of basic data of different Diopter groups.

| **Groups** | **n** | **age, year** | **Gender(M/F)** | **SE,D** | **IOP,mmHg** | **AL,mm** |
| --- | --- | --- | --- | --- | --- | --- |
| High myopia | 12 | 10.75±2.53 | 2/10 | -7.31±1.61 | 18.33±2.99 | 26.20±0.64 |
| Moderate myopia | 123 | 9.93±2.06 | 72/51 | -3.90±0.80 | 17.99±2.46 | 25.12±0.68 |
| Low myopia | 985 | 8.65±1.63 | 545/440 | -1.19±0.66 | 17.62±2.45 | 23.94±0.76 |
| Emmetropia | 2428 | 7.97±1.27 | 1285/1143 | 0.23±0.26 | 17.47±2.60 | 23.28±0.70 |
| Hyperopia | 2963 | 7.49±1.16 | 20/27 | 1.18±0.65 | 16.91±2.59 | 22.79±0.70 |
| statistic | | F=245.373 | $\chi^2 = 0.628$ | F=6044.100 | F=25.281 | F=201.312 |
| P |  | 0.000 |  | 0.000 | 0.000 | 0.000 |

### 2.2 Changes of AL and IOP in diopter groups

According to the five groups of diopter subjects, the respective differences among SE, IOP and AL are statistically significant between different groups (P < 0.001). Also, IOP and AL are positively related with diopter, while IOP is positively related with diopter and AL. The data of IOP and AL are higher in myopic eyes than in hyperopic eyes, and has a positive correlation with the degree of myopia, with ANOVA showing statistically significant differences between groups (P<0.01).(See Table 1)

### 2.3 Differences between different sex groups

The mean IOP in boys was (17.08±2.60) mmHg, the mean AL was (23.46±0.81) mm, and the spherical equivalent was (+0.34±1.18) D. The mean IOP in girls was (17.45±2.56)mmHg, the mean AL was (22.91±0.83)mm, and the spherical equivalent was (+0.37±1.21)D. The results of ANOVA showed that there were statistically significant differences in IOP data and AL data between male and female (P < 0.001). The average intraocular pressure of females was higher than that of males, which was consistent with the previous findings^[17-18]^.The average axial length of boys was higher than that of girls. There was no statistically significant difference between male and female equivalent spheres (P > 0.01), as shown inTable2 and Figure 1.

**Table 2.** Comparison of diopter, axial length and intraocular pressure of different genders.

| **Gender** | **n** | **Diopter (D)** | **AL(mm)** | **IOP(mmHg)** |
| --- | --- | --- | --- | --- |
| **Male** | **3470** | **0.34±1.18** | **23.46±0.81** | **17.08±2.60** |
| **Female** | **3041** | **0.37±1.21** | **22.91±0.83** | **17.45±2.56** |
| **F** |  | **1.426** | **719.845** | **34.510** |
| **P** |  | **0.233** | **0.000** | **0.000** |

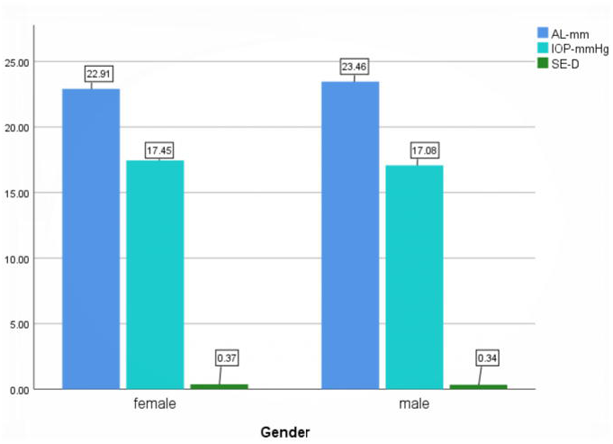

### 2.4 Differences between different age groups

Based on the distribution of the histogram (see Figure2), it can be initially found that IOP does not change significantly with age in children and adolescents aged 4-15 years, refraction progresses towards myopia with age, myopic refractive errors appear in children after the age of 10, and AL rises slowly with age and myopic refractive error.

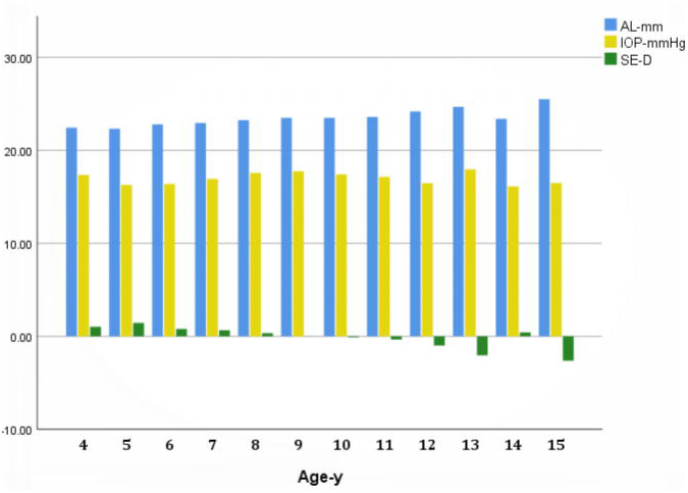

### 2.5 Correlation analysis of IOP, AL and SE

IOP showed a slight upward trend with the increase of AL, but showing a weak correlation (R =0.126, P < 0.001). There was no significant trend in IOP with increasing spherical equivalent, showing a weak correlation or no correlation (r=-0.116, P<0.001). AL was negatively correlated with the spherical equivalent. The higher the myopic diopter of axial myopia, the longer the axial length.(r=-0.632, P<0.001) (see Figure3)

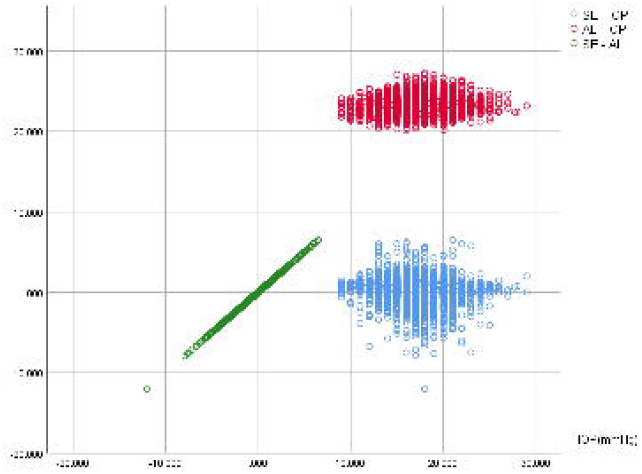

#### 2.5.1 Correlation between IOP, AL and SE in different diopter groups

##### 2.5.1.1 Hypermetropia group(n=2963)

There was a weak correlation between IOP and AL in hyperopic group (R =0.087, P < 0.01). IOP had no obvious change trend with the increase of spherical equivalent, showing weak or no correlation (r=-0.052, P < 0.01). The AL was negatively correlated with the SE (r=-0.400, P < 0.001), as shown in Figure 4.

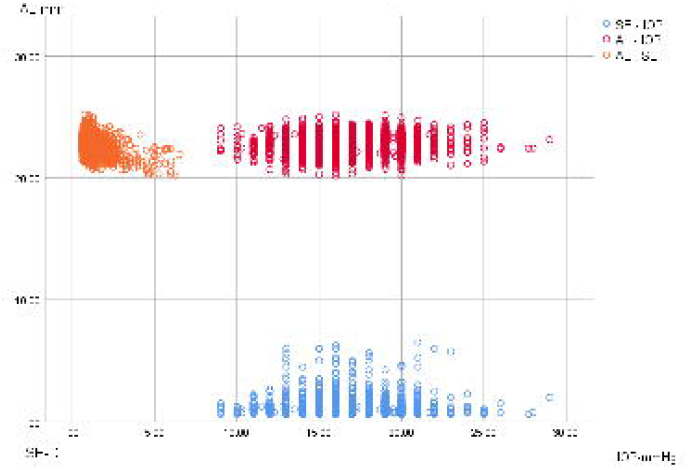

##### 2.5.1.2 emmetropic group(n=2428)

In the emmetropic group, there was a weak correlation between IOP and AL(r=0.072, P< 0.01). IOP had no obvious change trend with the increase of spherical equivalent, showing weak correlation(r=-0.064, P<0.01). There was a weak correlation between AL and SE (r=-0.123, P < 0.001). Different children’s height, weight and gender ^[19]^lead to different axial length, so it can be concluded that the AL of children’s eyes in the emmetropic group has no significant correlation with diopter. AL is closely related to height development in children and adolescents. It has been reported ^[20]^ that for every 1cm increase in the height of children and adolescents, the AL of persistent myopia will increase by 0.03mm. Some researchers found ^[21]^that subjects with higher BMI had higher diopters. (see Figure5)

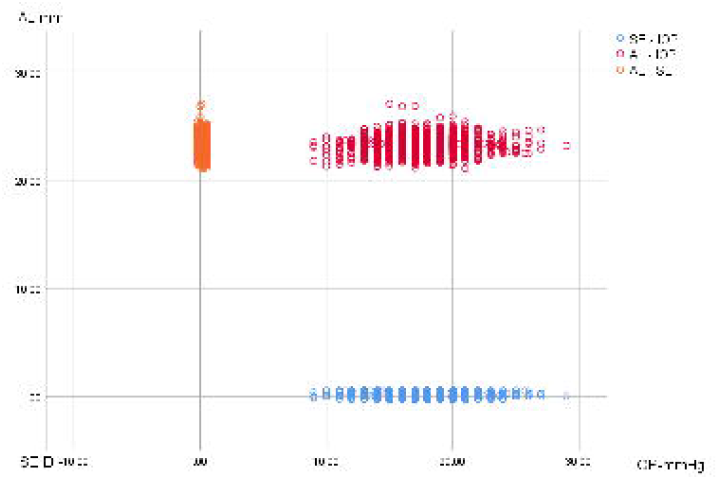

##### 2.5.1.3 Low-myopia group(n=985)

There was no correlation between IOP and AL in low-myopia group (R =0.044, P > 0.01). No significant change in IOP with the increase of SE (r=-0.009, P > 0.01) were found. There was a negative correlation between AL and SE, and the AL increased with the increase of myopic spherical equivalent (R =-0.407, P < 0.001).

##### 2.5.1.4 Medium and high myopia group(n=135)

IOP showed a slight upward trend with the increase of AL, and there was no linear correlation between IOP and AL (R =0.185, P > 0.01). There was no significant change in IOP with the increase of SE (r=-0.079, P > 0.01). There was a negative correlation between AL and SE, and the AL increased with the increase of myopic spherical equivalent (r=-0.549, P < 0.001).

## 3. Discussion

With the high incidence of the myopia in low age groups in China, the correlation between intraocular pressure, axial length and refraction has attracted more and more attention and discussion. As early as 1985, some foreign scholars found ^[22]^that there was a correlation between intraocular pressure and axial length in children’s growth and development. He found that children aged 1-3 years had the fastest eyeball development, and their axial length increased rapidly from 17mm to 22mm, accompanied by high intraocular pressure of 28mmHg in this period; After the age of 3, IOP gradually decreases, and the AL of children aged 3-15 grows slowly, about 1mm per year. People have further explored the correlation between the occurrence and development of myopia and IOP, and found that there are many positive links between myopia and IOP. Clinically, The intraocular pressure of myopic patients is higher than that of emmetropia patients^[1-3,23]^, and the intraocular pressure of high myopic groups is higher than that of medium and low degree ones, and the increase of diopter of axial myopia is accompanied by the growth of axial length. In this study, it was found that the mean IOP and AL of the hyperopia group were lower than those of the emmetropia group and the myopic group, and the IOP of the high myopia group was higher than that of the middle, low myopia group and the emmetropia group. The higher the diopter, the higher the IOP and ocular axis values were, and the difference between different diopters was statistically significant (P < 0.001), which was consistent with the previous research results^[24]^.Pearson correlation analysis of the data in this study showed that there was a positive correlation between AL and diopter in different diopter groups, which has long been the consensus of clinical experts. The increase of IOP in different diopter groups was weakly or not correlated with the increase of AL and diopter, which was consistent with previous studies at home and abroad^[25-28]^, indicating that IOP had no direct influence on the refractive state of children and adolescents, and it could not be proved that the change of IOP would have an impact on the occurrence and development of myopia in children and adolescents. At the same time, this study also found that the average intraocular pressure of girls was higher than that of boys (P < 0.01), which Costagliola et al. ^[29]^believed was related to the change of aqueous humor flow through trabecula caused by female hormone secretion. The average axial length of male eyes was higher than that of female eyes (P < 0.01), which might be related to the fact that male eyes were taller and heavier than female eyes at the same age.

Intraocular pressure is considered to be one of the factors related to myopia, though it is still under debate without reaching a definite conclusion. At present, the controversial points are whether intraocular pressure is related to the occurrence or development of myopia^[30]^, the sequence of occurrence between increased intraocular pressure and myopia, and whether the development of myopia can be effectively controlled by reducing intraocular pressure. Han et al. ^[31]^believe that myopia progresses faster in children with higher level of IOP than in children with lower level of IOP. Edwards and Goss^[32]^ found that intraocular pressure did not increase significantly before the occurrence of myopia, but increased to varying degrees with the occurrence of myopia. The mechanism of the increase of intraocular pressure caused by myopia is not clear. It may be related to the elongation of the scleral fiber wall caused by the increase of ocular axis, which reflects the increase of intraocular pressure^[33]^,It may also be related to the obstruction of aqueous humor circulation due to the structural changes of trabecular meshwork. An earlier study ^[34]^in monkeys found that elevated intraocular pressure in nearsightedness was associated with chronic over-regulation of focus. Rabbit experiments have confirmed^[35]^ that high intraocular pressure will lead to the growth of ocular axis, which may be related to the scleral wall thinning due to the increase of ocular axis, and the scleral wall thinning due to increased intraocular pressure^[36]^ or the decrease of choroid blood perfusion area, which causes choroid hypoxia and promotes the development of ocular axis. Based on this theory, if increased intraocular pressure causes the sclera wall to stretch, then some scholars believe that the use of IOP-lowering medication should slow the hypoxia of the choroid and thus delay the development of myopia. However, FDM models and LIM lens models in chicks confirmed^[37]^ that a significant reduction in IOP had little effect on the development of myopia. In another animal study^[38]^, tissue may be less likely to contract under the influence of a small drop in intraocular pressure. The study of Yan Hong et al. ^[39]^showed that the normal range of IOP had no correlation with diopter and axial length, so the local application of IOP drugs to control the development of myopia may not be effective. Some studies have found ^[40]^that reducing IOP can thick the choroid thickness, which may improve the blood circulation of the fundus, thus improving the fundus damage caused by high myopia. Others have found that reducing IOP can shorten the ocular axis, which may be caused by the thickening of the choroid thickness, but this does not prove that IOP is correlated with diopter.

## 4. Conclusion

This study suggests that there is no direct relationship between axial length and refraction or intraocular pressure, and intraocular pressure may not be a direct factor influencing the development of myopia. Some studies have found that there is a certain correlation between high myopic axis and IOP, which may be related to a certain intraocular pressure limit (threshold) that the eyeball can withstand. Grytz ^[41]^ and Markov^[42]^ et al. pointed out that the scleral tension in high myopic eyes increased, and when the intraocular pressure exceeded a certain range, the eyeball was more likely to expand and the posterior wall of the eye was thinner, thus promoting the growth of the ocular axis. Some studies have found ^[43]^through mechanical experiments that the IOP and axis of each normal eye match, the intraocular pressure within a certain range can be accepted by the eyeball has a safe range of axial length. However, when the axial growth exceeds the safe threshold, the eyeball may be slightly damaged due to physical compression, at which point myopic eyes may protect the eyeball by lowering the intraocular pressure. S M Lee et al. believed^[44]^ that the number of ametropia in children with anisometropia was not correlated with differences in axial length and intraocular pressure, and was more likely caused by differences in scleral structure determined by genes. At the same time, he observed the correlation between baseline IOP and myopia progression in Chinese children for 2 years, and found^[45]^ that there was basically no relationship between the two, while the lower IOP in progressive myopia may be related to the better compliance of the sclera. Nevertheless, we still need to pay close attention to the intraocular pressure in children and adolescents, especially in children with high myopia, because according to clinical observation, the intraocular pressure in children with myopia increases to varying degrees, and high intraocular pressure and high myopia are risk factors for the development of glaucoma in adolescents. Different studies have different conclusions, which may also be different from those obtained due to data errors caused by different methods of IOP measurement, different sample groups and age groups, ethnic differences and small sample size. At the same time, we should strengthen the detection of IOP in myopic patients, especially those with high myopia. For the myopia patients with high intraocular pressure and rapid increase in degree, if necessary, the screening of glaucoma should be excluded and the intraocular pressure should be reduced for treatment.

## Data Availability

All data produced in the present study are available upon reasonable request to the authors
All data produced in the present work are contained in the manuscript
All data produced are available online at

## Fund

Sichuan Provincial Department of Science and Technology, Research on Prevention and Control of Adolescent myopia by traditional Chinese Medicine, 2017SZ0023, Duan Junguo, Chengdu University of traditional Chinese Medicine, 20170101-20181231

## ETHICS STATEMENT

This study was approved by Medical Ethics Committee of Chengdu Ineye Hospital of Traditional Chinese Medicine (ID:2021-yh007)

## Acknowledgement

First of all, I would like to express my heartfelt thanks and high respect to my tutor Professor Duan Junguo for his care, help and guidance. At the same time, I would also like to express my heartfelt thanks to Dr. Jian Wenhuan, Dr. Yang Yanrong, Zhang Fuwen and other teachers and students for their kind help to me. In the process of writing papers, I met a lot of problems, always get Jian let teacher kindly care and guidance, make my paper Can be done fast and good, fu-wen zhang teacher with profound knowledge, rigorous doing scholarly research attitude, realistic working style and his quick thinking left a deep impression on me, I will never forget Mr. Zhang’s kind care and careful guidance. Finally, I would like to thank my family. Without your support, I would not be where I am today.

## References

[1] Quinn GE. Berlin JA, Young TL et al Association of intraocular pressure and myopia in children Ophthalmology. 1995. 102: 180∼185

[2] Edwards MH, Brown B (1993) Intraocular pressure in a selected sample of myopic and nonmyopic Chinese children. Optom Vis Sci 70: 15–17. pmid:8430003

[3] Xue Yushun, Li Yuqin, Shi Rui, Wang Ke. Correlation analysis of diopter with axial length and intraocular pressure in adolescent myopia [J]. International Journal of Ophthalmology,2008(09):1847–1849.

[4] Weih LM, Mukesh BN, McCarty CA, et al. Asssociation of demographic, familial, medical, and ocular factors with intraocular pressure. Arch Ophthalmol 2001;119:875–80. PubMedWeb of ScienceGoogle Scholar

[5] Quinn GE, Berlin JA, Young TL, et al. Association of intraocular pressure and myopia in children. Ophthalmology 1995;102:180–5. PubMedWeb of ScienceGoogle Scholar

[6] Giloyan A, Harutyunyan T, Petrosyan V. Risk Factors for Developing Myopia among Schoolchildren in Yerevan and Gegharkunik Province, Armenia. Ophthalmic Epidemiol 2017;24(2):97–103

[7] Leydolt C, Findl O. Drexler W. Effects of change in intraocular pressure on axial eye length and lens position[J]. Eye, 2008, 22 (5):657—661.

[8] Vohra SB, Good PA. Altered globe dimensions of axial myopia as risk factors for penetrating ocular injury during peribulbar anaesthesia. Brl Anaesthesiol 2000;85(2):242—245

[9] Francis BA, Wang M, Lei H, LT Du, MincMer DS, Green RL, etal. Changes in axial length following trabeculectomy and glaucoma drainage device surgery[J]. BrJOphthalmol, 2005, 89(1):17—20.

[10] Puell Marin MC, Romero Martin M, Dominguez Carmona M. JAmOptomAssoc 1997 68:657

[11] Manny RE, Deng L, Crossnoe C, et al. IOP, myopic progression and axial length in a COMET subgroup. Optometry Vis Sci 2008;85(2):97–105

[12] Masayuki H, Fumitaka H, Akio O, Yasuhiko H, Yasuo K. Changes in choroidal thickness and optical axial length accompanying intraocular pressure increase[J]. Jpn J Ophathalmol, 2012, 56(6); 564–568.

[13] Yu Teng-Chieh, Wu Tzu-En, Wang Yuan-Shen et al. A STROBE-compliant case-control study: Effects of cumulative doses of topical atropine on intraocular pressure and myopia progression. [J]. Medicine (Baltimore), 2020, 99: e22745.

[14] Al-Obaida Ibrahim, Al Owaifeer Adi Mohammed, Ahmad Khabir et al. The relationship between axial length, age and intraocular pressure in children with primary congenital glaucoma. [J]. Sci Rep, 2020, 10: 17821.

[15] Read SA, Collins MJ, Iskander DR. Diurnal variation of axial length, intraocular pressure, and anterior eye biometrics. Invest Ophthalmol Vis Sci 2008;49(7):2911–2918

[16] Fiedorowicz Michal, Welniak-Kaminska Marlena, Swiatkiewicz Maciej et al. Changes of Ocular Dimensions as a Marker of Disease Progression in a Murine Model of Pigmentary Glaucoma. [J]. Front Pharmacol, 2020, 11: 573238.

[17] Li S, Li SM, Wang XL, Kang MT, Liu LR, Li H, Wei SF, Ran AR, Zhan S, Thomas R, Wang N; Anyang Childhood Eye Study Group. Distribution and associations of intraocular pressure in 7- and 12-year-old Chinese children: The Anyang Childhood Eye Study. PLoS One. 2017 Aug 17;12(8):e0181922. doi: 10.1371/journal.pone.0181922. PMID: 28817606; PMCID: PMC5560658.

[18] Li SM, Iribarren R, Li H, Kang MT, Liu L, Wei SF, Stell WK, Martin G, Wang N. Intraocular pressure and myopia progression in Chinese children: the Anyang Childhood Eye Study. Br J Ophthalmol. 2019 Mar;103(3):349–354. doi: 10.1136/bjophthalmol-2017-311831. Epub 2018 Jun 1. PMID: 29858181.

[19] Selović A, Juresa V, Ivankovic D, et al. Relationship between axial length of the emmetropic eye and the age, body height, and body weight of schoolchildren. Am J Hum Biol. 2005;17(2):173–177.

[20] Kearney S, Strang NC, Cagnolati B, et al. Change in body height, axial length and refractive status over a four-year period in caucasian children and young adults. J Optom. 2020;13(2):128–136.

[21] Roy A, Kar M, Mandal D, et al. Variation of Axial Ocular Dimensions with Age, Sex, Height, BMI-and Their Relation to Refractive Status. J Clin Diagn Res. 2015;9(1):AC01–AC4.

[22] Curtin, BJ. The myopias-basic science and clinical management. Philadelphia:Harper &Row, Publishers, 1985:3–59.

[23] EdW8rds MH. &uwn B Ophthahlc PhysIol opt 1996 16:243

[24] Manny RE, Mitchell GL, Cotter SA, et al. Intraocular pressure, ethnicity, and refractive error. Optom Vis Sci, 2011, 88(12): 1445–1453. DOI: 10.1097/OPX.0b013e318230f559.

[25] Urban B, Bakunowicz-Lazarczyk A. Cisnienie wewnatrzgalkowe u dzieci i mlodziezy z krótkowzrocznoscia [Intraocular pressure in children and adolescents with myopia]. Klin Oczna. 2010;112(10-12):304–6. Polish. PMID: 21473082.

[26] Lee AJ, Saw SM, Gazzard G, Cheng A, Tan DT. Intraocular pressure associations with refractive error and axial length in children. Br J Ophthalmol. 2004 Jan;88(1):5–7. doi: 10.1136/bjo.88.1.5. PMID: 14693759; PMCID: PMC1771921.

[27] Li SM, Iribarren R, Li H, Kang MT, Liu L, Wei SF, Stell WK, Martin G, Wang N. Intraocular pressure and myopia progression in Chinese children: the Anyang Childhood Eye Study. Br J Ophthalmol. 2019 Mar;103(3):349–354. doi: 10.1136/bjophthalmol-2017-311831. Epub 2018 Jun 1. PMID: 29858181.

[28] Yan Hong, Yan Ling, Zhang Shuang, et al. Intraocular pressure and children’s refractive errors with ocular axial length relationship [J]. Chinese journal of practical ophthalmology, 2004, 22 (11) : 927–928. The DOI: 10.3760/cma.J.iSSN.1006-4443.2004.11.027.

[29] Costagliola CIRO, Trapanese Antonio, Pagano MONICA. Intraocular Pressure in a Healthy Population: A Survey of 751 Subjects[J]. Optometry & Vision Science, 1990. 204–206.

[30] Lee Kwanghyun, Yang Heon, Kim Joo Yeon, Seong Gong Je, Kim Chan Yun, Bae Hyoung Won. Risk Factors Associated with Structural Progression in Normal-Tension Glaucoma: Intraocular Pressure, Systemic Blood Pressure, and Myopia.[J]. Investigative ophthalmology & visual science, 2020, 61(8).

[31] Han X, Yang T, Zhang J, et al. Longitudinal changes in intraocular pressure and association with systemic factors and refractive error: Lingtou Eye Cohort Study. BMJ Open, 2018, 8(2): e019416. DOI: 10.1136/bmjopen-2017-019416

[32] GossDA, CaHeylw o|m VB sci 1999 76:286

[33] Greene PR. Mechanical consideration in myopia. In Grosvenor T, Flam Mc (Eds), Refractive nomalies :Res Clin Appl, 1991; 287–300.

[34] Young FA. The development and control of myopia in human and sub— human primates. Contacto. 1975, 19(6):16—31.

[35] Zhang Jian-hua, Wang Yu-dong, Jin Jia-yan, Shi Rong-xian, Li Bin, Chao Xiao-rui. Effects of intraocular pressure on axial length of eye [J]. Advances in ophthalmology, 2013, 33(09):826–829.

[36] Tomlinson A, Phillips CI. Applanation tension and axial length of the eyeball. British Journal of Ophthalmology, 1970

[37] Close K. L. Schmid, M. Abbott, M. Humphries, K. Pyne, C. F. WildsoetTimolol lowers intraocular pressure but does not inhibit the development of experimental myopia in chick Experimental Eye Research, 70 (5) (2000), pp. 659–666

[38] Goldberg LA, Rucker FJ. Opposing effects of atropine and timolol on the color and luminance emmetropization mechanisms in chicks. Vision Res. 2016 May;122:1–11. doi: 10.1016/j.visres.2016.03.001. Epub 2016 Mar 19. PMID: 26971621; PMCID: PMC4861675.

[39] Yan Hong, YAN Ling, ZHANG Shuang, Yan Ming. Relationship between intraocular pressure and refractive error and axial length in children [J]. Chinese Journal of Practical Ophthalmology, 2004(11):927–928.

[40] Li Min, CHENG Huiqin, YUAN Ying, WANG Jia, KE Bilian. Relationship between choroid thickness and diopter and intraocular pressure in myopia [J]. Chin J optometry & vision science, 2014, 16(06):350–353.

[41] Grytz R, Siegwart JJT. Changing material properties of the tree shrew sclera during minus lens compensation and recovery[J]. Investig.Ophthalmol.Vis.Sci, 2015(56):2065–2078.

[42] Markov PP, Eliasy A, Pijanka JK, et al. Bulk changes in posterior scleral col-lagen microstructure in human high myopia[J]. Mol.Vis, 2018(24):818–833.

[43] Annival Abdukrim, Wang Wei, Ni Wei. The relationship between intraocular pressure and ocular axis [J]. Acta Psychologica Sinica, 2012, (3):52–54. (in Chinese) DOI:10.3969/j.issn.1007-8231.2012.03.047.

[44] Lee SM, Edwards MH. Intraocular pressure in anisometropic children. Optom Vis Sci. 2000 Dec;77(12):675–9. doi: 10.1097/00006324-200012000-00015. PMID: 11147738.

[45] Li SM, Iribarren R, Li H, et al. Intraocular pressure and myopia progression in Chinese children: the Anyang Childhood Eye Study. Br J Ophthalmol. 2019;103(3):349–354.

